# Rapid and sensitive detection of SARS-CoV-2 antibodies by biolayer interferometry

**DOI:** 10.1101/2020.07.17.20156281

**Authors:** John V. Dzimianski, Nicholas Lorig-Roach, Sara M. O’Rourke, David L. Alexander, Jacqueline M. Kimmey, Rebecca M. DuBois

## Abstract

Serological testing to evaluate antigen-specific antibodies in plasma is generally performed by rapid lateral flow test strips that lack quantitative results or by high complexity immunoassays that are time- and labor-intensive but provide quantitative results. Here, we describe a novel application of biolayer interferometry for the rapid detection of antigen-specific antibody levels in plasma samples, and demonstrate its utility for quantification of SARS-CoV-2 antibodies. Our biolayer interferometry immunosorbent assay (BLI-ISA) utilizes single-use biosensors in an automated “dip-and-read” format, providing real-time optical measurements of antigen loading, plasma antibody binding, and antibody isotype detection. Complete quantitative results are obtained in less than 20 minutes. BLI-ISA meets or exceeds the performance of high complexity methods such as Enzyme-Linked Immunosorbent Assay (ELISA) and Chemiluminescent Immunoassay. Importantly, our method can be immediately implemented on existing BLI platforms for urgent COVID-19 studies, such as serosurveillance and the evaluation of vaccine candidates. In a broader sense, BLI-ISA can be developed as a novel diagnostic platform to evaluate antibodies and other biomolecules in clinical specimens.

## Introduction

In December 2019, a novel coronavirus emerged in Wuhan, China, causing severe respiratory disease with initial reported fatality rates of 2-3% ^1^. In the ensuing months the virus became established internationally through travel and community transmission, leading to the declaration of a pandemic by the WHO on March 11, 2020 ^2^. Officially named severe acute respiratory coronavirus 2 (SARS-CoV-2) by the International Committee on Taxonomy of Viruses due to its phylogenetic relatedness to SARS and SARS-like coronaviruses ^3^, the virus causes <u>co</u>rona<u>vi</u>rus <u>d</u>isease 20<u>19</u> (COVID-19). As of July 16, 2020, over 13 million cases and over 580 thousand deaths have been reported due to COVID-19, and the disease continues to be a source of economic and societal strain. Efficient and accurate testing is critical to understand the full breadth of impact and to developing countermeasures to limit future infections.

Detection of SARS-CoV-2 infection relies predominantly on two approaches: nucleic acid testing, which detects viral RNA, and serological testing, which detects antibodies elicited against SARS-CoV2 antigens. Nucleic acid testing methods were quickly developed after the release of the virus genome ^4-6^ and serve as the primary, definitive diagnostic tool for active cases of COVID-19. However, due to limitations in nucleic acid testing availability and the occurrence of mild or asymptomatic infections, many cases of COVID-19 are not diagnosed. Thus, serological testing, which detects antibodies elicited by SARS-CoV-2 antigens, have become key to assessing the true extent of SARS-CoV-2 spread within the population ^7^.

Serological studies have shown that antibodies develop over several weeks following infection with SARS-CoV-2, and that antibody levels can vary significantly between individuals ^8-11^. Accurate serological testing is crucial to develop countermeasures against SARS-CoV-2 infection, including the identification and evaluation of donors for convalescent plasma therapy and the development of a SARS-CoV-2 vaccine.

Serological testing methods for SARS-CoV-2 predominantly use the virus nucleocapsid protein, the spike glycoprotein, or fragments thereof such as the spike receptor binding domain (RBD), to probe for antibodies. Methods that utilize the spike, and the RBD in particular, have been shown to correlate with SARS-CoV-2 neutralization assays ^8,10,12-17^. Serological tests are also distinguished on whether they detect total antibodies, IgG, IgM, or both IgG and IgM. Current tests that have been developed include the Lateral Flow Immunoassay (LFIA), Enzyme-Linked Immunosorbent Assay (ELISA), Immunofluorescent Assay (IFA), and Chemiluminescent Immunoassay (CLIA). LFIA tests present the most rapid turnaround and can be performed with minimal training, with test strip bands visualized in 15-20 minutes, making it useful as a point-of-care test ^18^. The output, however, is generally a positive/negative binary outcome, and the tests have had a mixed performance in terms of sensitivity ^19,20^. ELISA, IFA, and CLIA tests are high complexity laboratory tests that are generally robust in terms of the sensitivity and specificity ^20-22^. In addition, these methods provide a quantitative measure of antibody responses that can distinguish between strong and weak responses. However, ELISA, IFA, and CLIA are time-intensive processes requiring 1-5 hours, with significant incubation times and washing steps that are performed manually or require automated fluidic platforms. Due to these drawbacks, the development of alternate serological testing methods that are simple, rapid, and quantitative would be advantageous for many applications.

Here, we describe a novel method for measuring antigen-specific antibodies in blood plasma utilizing biolayer interferometry (BLI). BLI is a fiber optics-based biophysical technique designed to measure the affinity between biological molecules. White light is shone down a fiber optic biosensor and the interference between light reflected off two layers—a reference layer and a biological layer—is measured ^23^. Binding of molecules to the biosensor surface results in a real-time signal due to the shift in the wavelength of the reflected light. While historically used to precisely measure the kinetics of binding between purified biological molecules, BLI has also been adapted to quantify a target biological molecule in more complex fluids, such as proteins in cell growth media ^24,25^ and biomolecules in clinical specimens ^26-31^. We developed a novel application for this technology, termed biolayer interferometry immunosorbent assay (BLI-ISA), for the rapid and quantitative measurement of SARS-CoV-2 antibodies in plasma. BLI-ISA advantages include a simple “dip-and-read” format that is free of fluidics, provides real-time measurements of both total antibody levels and specific antibody isotypes in the same assay, and can detect weakly seropositive samples. Importantly, BLI-ISA can be completed in less than 20 minutes and provides quantitative information on antibody levels. This method can be immediately implemented for urgent SARS-CoV-2 pandemic studies, including evaluation of antibody responses to natural infection and to candidate vaccines, as well as serosurveillance studies. In a broader sense, BLI-ISA can be adapted and multiplexed, allowing for measurement of antibodies against multiple antigens, detection of multiple antibody isotypes, and quantitation of other clinically relevant biological molecules.

## Results

### Validation of test samples by ELISA

For the development of serological testing, we obtained 37 plasma or serum samples to serve as our test set for experimental design. This included 10 commercially available convalescent plasma samples from donors who had recovered from COVID-19, each of which had previously tested positive by a CoV2T CLIA test, with CoV2T scores ranging from 8 to 440 (Table 1). These ten samples served as our presumed seropositive (SP) group. 27 plasma and serum samples collected prior to the COVID-19 pandemic formed the presumed seronegative (SN) group.

**Table 1.**
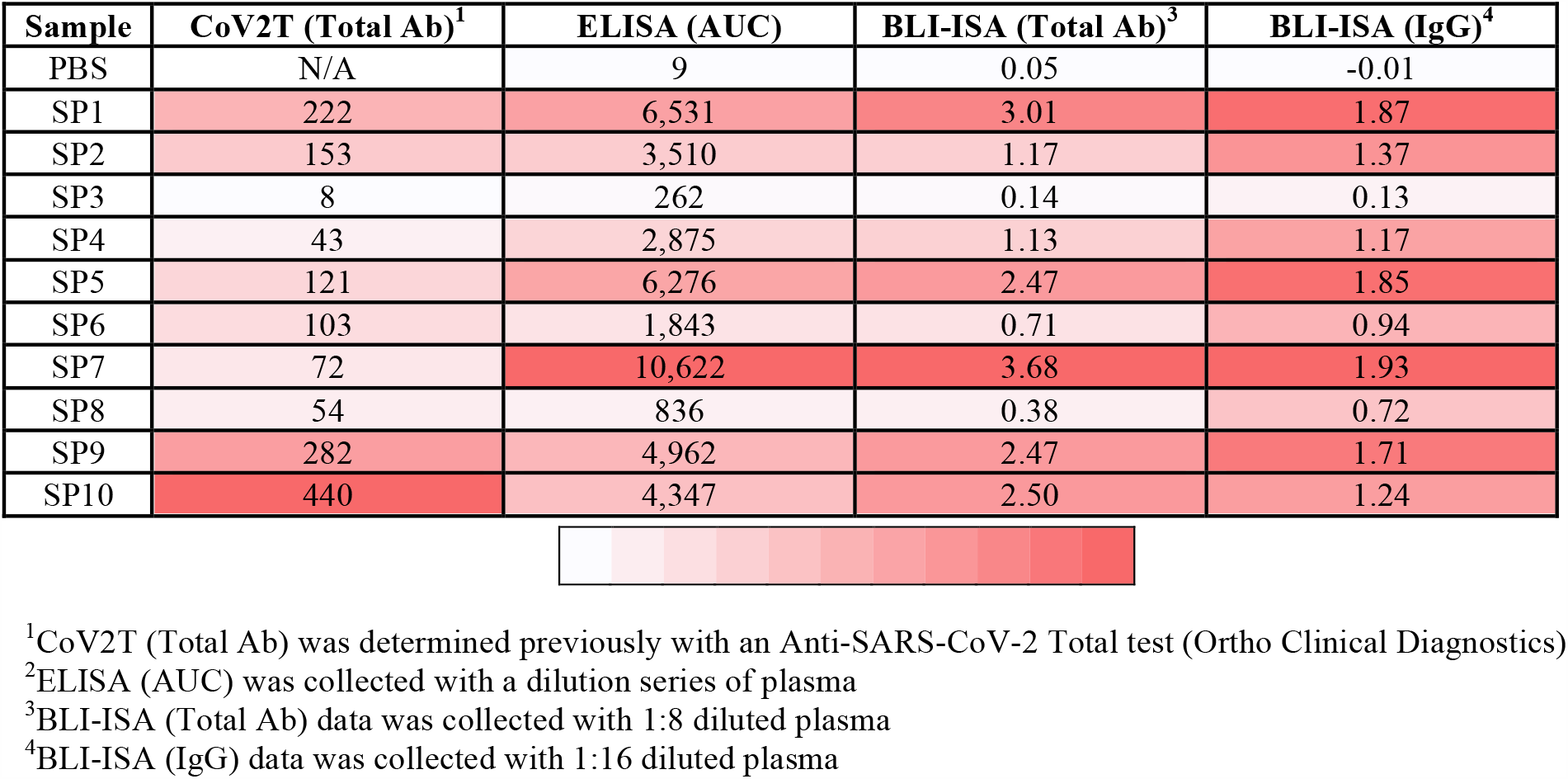
Comparison of antibody reactivity assays with SARS-CoV-2 RBD antigen.

To confirm the status of the samples and establish the expected outcomes for future study, we adapted a published ELISA protocol to measure IgG antibody reactivity towards recombinant SARS-CoV-2 spike RBD (Fig 1, Supplementary Figure 1)^12^. All samples were evaluated at 1:50 dilution in biological duplicates with RBD-coated plates as well as control uncoated plates. The majority of pre-pandemic (presumed seronegative) samples did not react to RBD-coated plates, resulting in a signal that was indistinguishable from the background (mean SN OD_490_ = 0.227). As expected, the presumed positive samples showed a robust signal in ELISA that was substantially higher than the presumed negatives and background in the absence of antigen (Fig 1a). Intriguingly, however, there were two notable exceptions to these trends. The presumed positive sample SP3 failed to exceed its own background and was indistinguishable from some of the presumed negatives. In contrast, SN22 showed a signal above background that, while weaker than most of the presumed positives, appeared to be more robust than SP3.

**Fig. 1.**
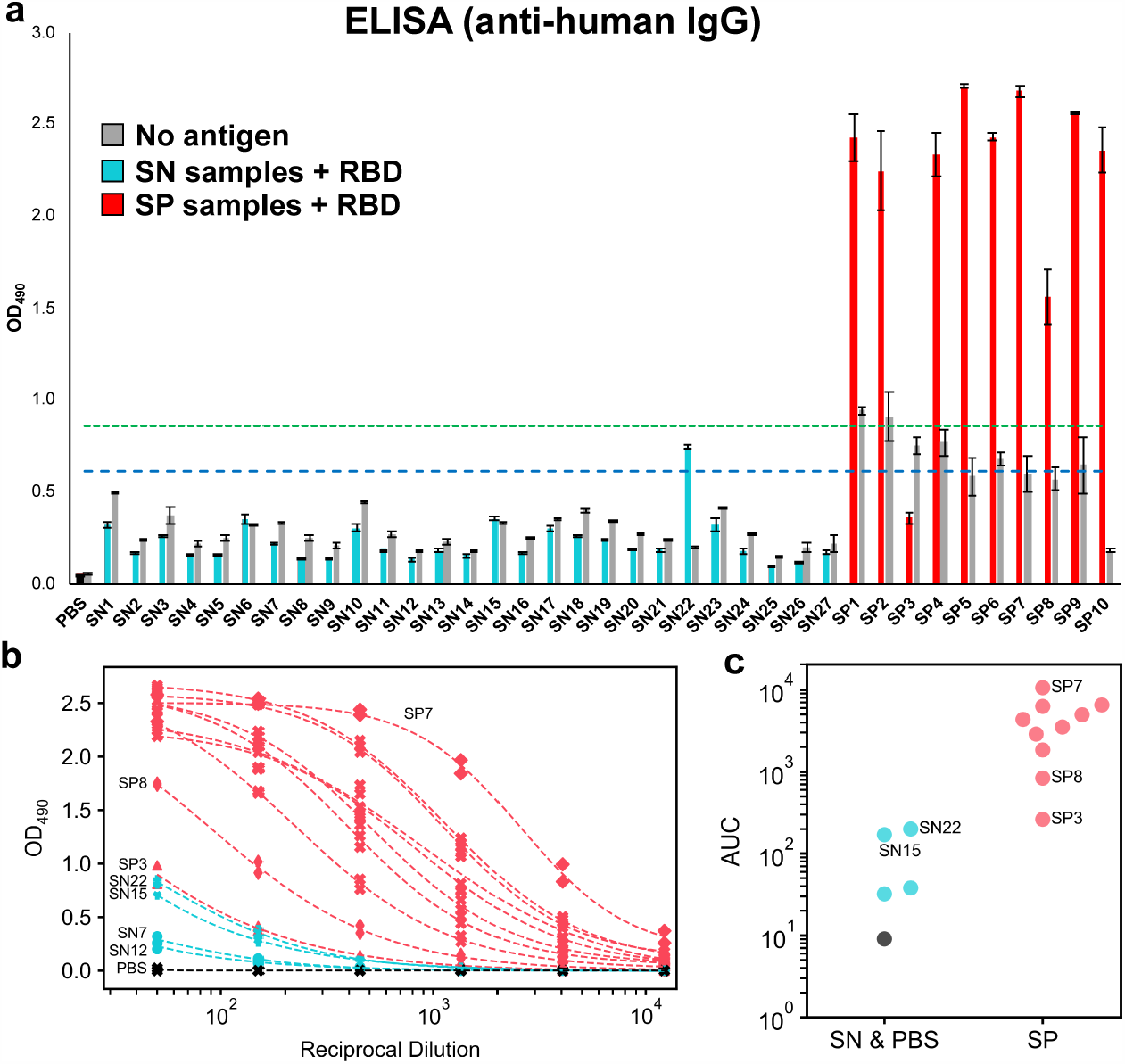
ELISA evaluation of SARS-CoV-2 spike RBD reactivity of pre-pandemic and convalescent plasma. **a** Single-dilution ELISA to evaluate the presence of RBD-reactive human IgG in pre-pandemic seronegative (SN, cyan) and convalescent seropositive (SP, red) samples compared to no-antigen controls (grey). The assays were performed with plasma at a 1:50 dilution. Samples were evaluated in biological duplicates and error bars represent one standard deviation from the mean. Blue and green dashed lines represent the mean of seronegative samples plus 3 and 5 standard deviations, respectively. **b** Dilution series ELISA was performed to quantitate RBD-reactive human IgG in plasma. Samples were evaluated in biological duplicates. Dashed curves represent fit lines from a four-parameter logistic regression applied over each series. **c** Data from **b** plotted as area-under-the-curve (AUC).

We subsequently performed dilution series ELISA and area-under-the-curve (AUC) calculations for all SP samples and a subset of SN samples (SN7, SN12, SN15, and SN22) chosen to represent the diversity of signals observed in the single-dilution ELISA (Fig 1b, 1c, Table 1). The dilution series ELISA clarified the differences between highly reactive samples, revealing SP7 as having the most robust anti-CoV-2 spike RBD IgG levels, while SP8 showed moderate levels. In contrast, SP3 overlapped with SN22 and was only slightly more reactive than SN15.

### Design and optimization of antibody detection by BLI-ISA

Having established the range of expected reactivity of the samples for SARS-CoV-2 spike RBD by ELISA, we then developed the BLI-ISA method. We developed this method with the goal of a simple, rapid (<20 minutes), and quantitative method to measure antigen-specific antibodies in plasma (Fig. 2). The simple “dip-and-read” format allows for antigen and antibody samples to be plated into a 96- or 384-well plate (Supplementary Figure 2). This plate is then loaded along with single-use biosensors into the BLI instrument, which dips the biosensors into designated plate wells to perform each step. Most notably, the BLI technology enables real-time measurements throughout the entire experiment (Supplementary Figure 3). The signal from the antigen loading step serves as a quality control measure and ensures even antigen loading onto each biosensor surface. In addition, two antibody-binding steps can be evaluated by our method; (1) a Total Antibody Binding signal is measured when the antigen-coated biosensors are dipped into plasma samples and (2) a Detection Antibody Binding signal is measured when the biosensors are subsequently dipped into an anti-human IgG secondary antibody reagent.

**Fig. 2.**
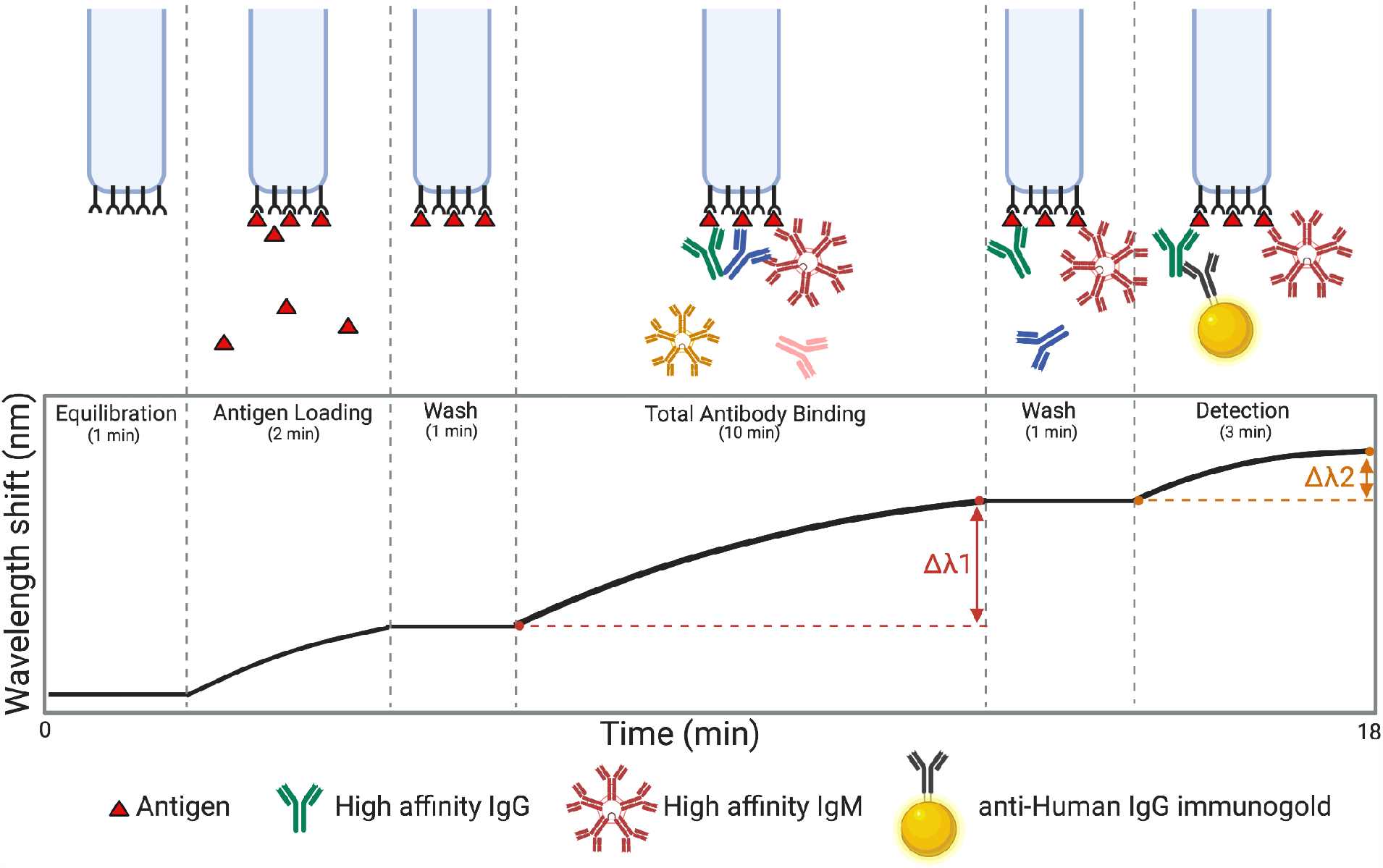
Overview of the BLI-ISA experiment. To begin, a tray of fiber optic biosensors and a 96- or 384-well plate of samples are placed into the Octet BLI instrument (Supplementary Figure 2), and the assay program is run. Throughout the experiment, real-time measurements are recorded as the change in the wavelength of reflected light returning from the biosensor surface. First, biosensors are equilibrated by dipping into wells containing BLI assay buffer. In the antigen loading step, biosensors are dipped into wells containing tagged antigen (e.g. streptavidin SA biosensors dipped into biotinylated antigen). After a wash, antigen-loaded biosensors are placed into diluted plasma, and a Total Antibody Binding signal is measured. After another wash, the antigen-antibody-coated biosensors are dipped into wells containing isotype-specific binding reagents (e.g. colloidal gold-conjugated anti-human IgG), and a Detection signal is measured. Created in BioRender.com.

Several features required optimization including antigen loading, assay buffers, plasma dilution factors, secondary antibody detection reagents, and times at each assay step. First, we evaluated the loading stability of three recombinant SARS-CoV-2 spike antigens onto BLI biosensor tips: the His-tagged RBD (RBD-His) used for ELISA ^12^, an Avi-tagged RBD with a single biotin modification on the Avi-Tag (RBD-biotin), and the His-tagged prefusion-stabilized spike trimer (prefusion Spike-His) ^32,33^(Supplementary Figure 1). All three constructs exhibited sufficient loading onto their respective anti-penta-His (HIS1K) or (anti-biotin) streptavidin (SA) biosensors. However, loading of RBD-His demonstrated considerable downward baseline drift compared to RBD-biotin with SA biosensors (Supplementary Figure 4). Interestingly, this downward drift was not observed by the prefusion Spike-His, likely due to the stronger anchoring by the three His-tags in this trimeric form. To enable comparison of our BLI-ISA results with the CoV2T CLIA and ELISA, which use the RBD antigen, we further optimized our method using RBD-biotin loaded onto SA biosensors. During buffer optimization, we found that addition of 20-25% ChonBlock, a blocking agent, to the plasma samples reduced background signal from SN samples without affecting SP sample signals (Supplementary Figure 5). Finally, we developed a novel method to quantify the amount of antibody bound to the antigen-coated biosensor. Whereas anti-human antibodies alone or conjugated to enzymes did not yield a signal (data not shown), we discovered that colloidal gold-conjugated anti-human antibody reagents were large enough to give a significant signal in the Detection step (Fig 2 and Supplementary Figure 3).

To confirm the specificity of BLI-ISA, we tested antigen binding with commercially available rabbit antibodies that were raised against the spike proteins of either HCoV-HKU1 or SARS-CoV-2 (Supplementary Figure 6). Consistent with previous studies showing no cross-reactivity of antibodies against HCoV-HKU1 toward the SARS-CoV-2 spike RBD ^8,12^, polyclonal antibodies against HCoV-HKU1 spike showed a negligible Total Antibody Binding signal, whereas both polyclonal and monoclonal rabbit SARS-CoV-2-targeting antibodies resulted in a significantly higher Total Antibody Binding signal. As an additional check on the specificity, we evaluated the signals by the anti-human IgG secondary antibody in the Detection step. As a positive control, we used the human monoclonal antibody CR3022, which targets the SARS-CoV-1 RBD and has known cross-reactivity with the SARS-CoV-2 RBD ^34-36^. The anti-human IgG secondary antibody exhibited robust binding for CR3022 but only weak association with the rabbit antibodies, indicating a specific interaction.

Finally, we identified a 1:8 plasma dilution as an optimal screening dilution for BLI-ISA to minimize assay time and sample volume, while maximizing the dynamic range during the Total Antibody Binding step (Fig 3). Our assay utilizes a 10-minute Total Antibody Binding step with plasma, however if sample volume is limited, a higher dilution factor could be used, and this could be compensated for with a longer Total Antibody Binding step to achieve the same signal. Similarly, we identified a 1:10 secondary antibody dilution as an optimal dilution to minimize assay time and maximize the signal during the Detection step (data not shown).

**Fig. 3.**
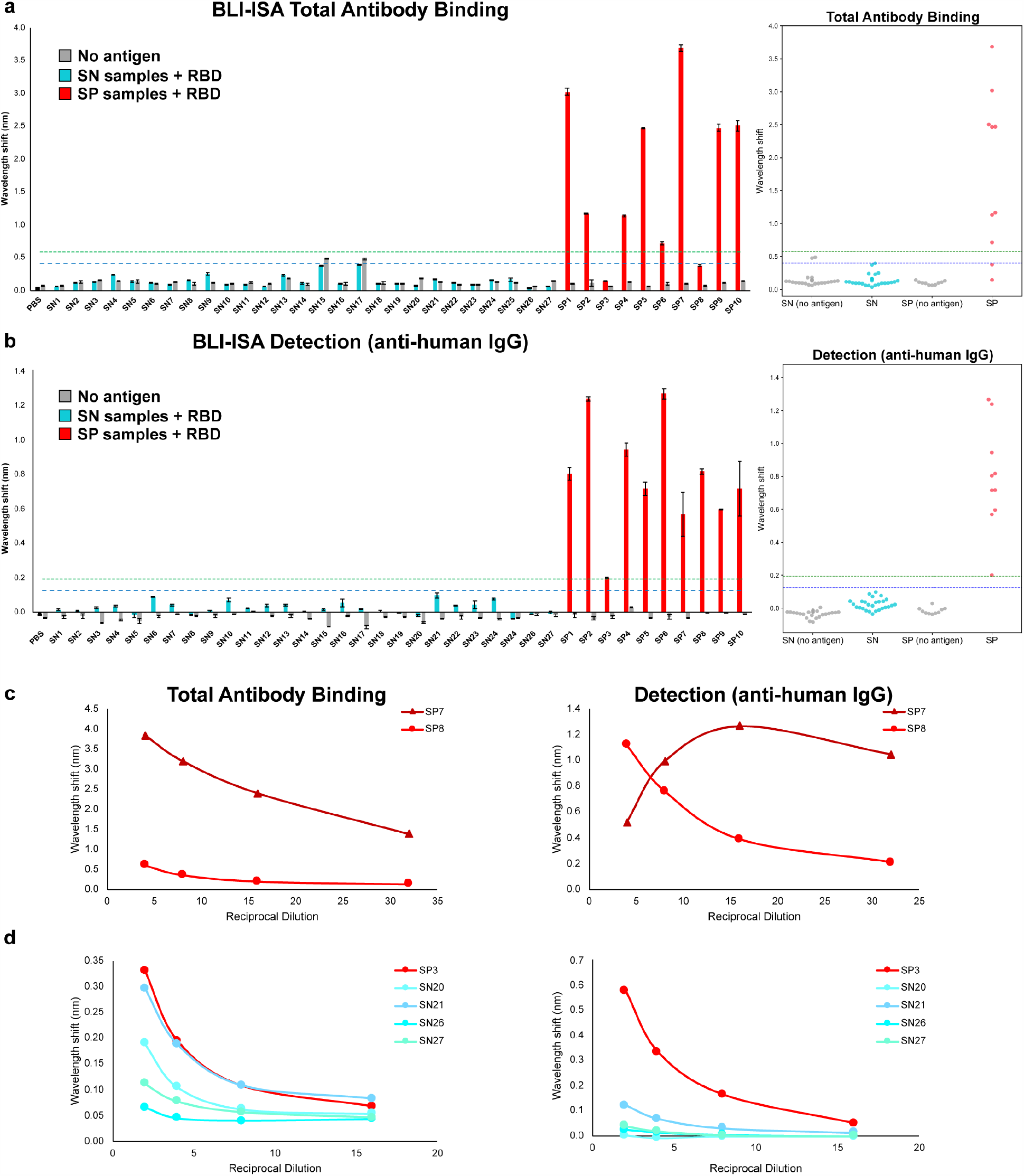
BLI-ISA evaluation of SARS-CoV-2 spike RBD reactivity of pre-pandemic and convalescent plasma. **a-b** Single-dilution BLI-ISA to evaluate the presence of RBD-reactive human antibodies in the pre-pandemic seronegative (SN, cyan) and convalescent seropositive (SP, red) samples compared to no-antigen controls (grey). The assays were performed with plasma at a 1:8 dilution. Bars and dots represent the mean of biological duplicates, and error bars represent one standard deviation from the mean. Blue and green dashed lines represent the mean of seronegative samples plus 3 and 5 standard deviations, respectively. **a** The Total Antibody Binding signal is measured when RBD-biotin-loaded SA biosensors are dipped into plasma samples. **b** The Detection signal is measured when RBD-biotin-loaded SA biosensors that had been dipped into plasma are subsequently dipped into colloidal gold-conjugated anti-human IgG. **c** Dilution series BLI-ISA from representative strong (SP7) and moderate (SP8) seropositive samples. **d** Dilution series BLI-ISA from the weakest seropositive sample (SP3) compared to seronegative plasma samples.

Importantly, the real-time measurements of the BLI-ISA allow this assay flexibility which, to our knowledge, is not practical with any other currently available serological assays.

### Measurement of plasma antibody levels by BLI-ISA

With the method optimized, we then performed BLI-ISA on our test set of 37 plasma samples (Fig 3). All samples were evaluated at 1:8 dilution in biological duplicates with RBD-biotin-coated biosensors as well as control “no antigen” biosensors. In contrast to other serological tests, which detect either total antibodies or isotype-specific antibodies, BLI-ISA gives signals in real-time as plasma antibodies bind, thus allowing for measurements at this Total Antibody Binding step, as well as measurements at the Detection step after addition of anti-human IgG secondary antibody binding. As a result, we report the results of these two steps separately as the overall change in signal at each step (Fig 2).

Data from the Total Antibody Binding step revealed that the strongest SP samples (8 of the 10 samples) could be easily distinguished from the SN samples (Fig 3a). Significantly, the trends obtained with this single dilution in BLI-ISA generally align with the AUC values obtained by dilution-series ELISA (Table 1), revealing single-dilution BLI-ISA as a rapid method to identify and differentiate antibody levels of strongly SP samples. Using a cutoff of the mean of seronegative samples plus three standard deviations (mean SN signal = 0.140; standard deviation = 0.088) unambiguously identifies 8 of the 10 SP samples as positive. SP3 and SP8 do not have sufficient RBD-specific antibody levels to test positive in this step. Two seronegatives (SN15 and SN17) are slightly positive, likely due to background signal intrinsic to these specific samples given that a similar signal is observed in the absence of antigen. Additionally, using a more conservative cutoff of the SN mean plus five standard deviations removes this ambiguity without impacting the remaining SP samples.

To specifically measure anti-RBD IgG levels, the sensors are then dipped in wells containing colloidal gold conjugated anti-human IgG. Data from this Detection step distinguishes between all 27 SN samples and 10 SP samples (Fig 3b). Here, all SP samples are positive, even with the more conservative cutoff of the SN mean plus five standard deviations. Notably, SP8 is clearly positive, consistent with ELISA results (Fig 1a), while SP3 is weakly positive. All the SN samples are below both cutoffs, and we no longer observe nonspecific background positivity (“no antigen” wells do not approach detection threshold).

Interestingly, the signals from the strong SP samples in the Detection step do not strongly correlate with their signals in the Total Antibody Binding step. This suggests a potential saturation effect, where the large number of antibodies bound during the Total Antibody Binding step saturates the biosensor surface and curtails the signal during the Detection step with anti-human IgG. To confirm this, we performed BLI-ISA with a dilution series ranging from 1:4 to 1:32 using SP7 and SP8, which respectively gave the strongest and second weakest signals in the Total Antibody Binding step, but relatively similar signals in the detection step (Fig 3c). As expected, the Total Antibody Binding step showed a predictable correlation between strength of signal and sample dilution for both SP7 and SP8. In contrast, while this trend is also present for SP8 in the Detection step, there is actually a lower signal for SP7 at the highest concentration of plasma, followed by a rebound before it begins to follow a normal pattern. Thus, there appears to be a maximum combined signal from the antibodies in both steps, limiting the anti-human IgG antibody binding signal to a qualitative role confirming the presence of antibodies in samples at the 1:8 dilution. To overcome this limitation, we evaluated all ten SP samples at a 1:16 dilution in biological duplicates and found that the signals in the Detection step now align with the respective signals in the Total Antibody Binding step as well as the AUC values obtained by dilution series ELISA (Table 1).

Given the observed boost in signal in SP8 at a 1:4 dilution, we investigated whether a follow-up assay with a higher proportion of plasma could boost the signal in the case of very weak seropositives, such as SP3. We performed a dilution series ranging from 1:2 to 1:16 with SP3 and the seronegatives SN20, SN21, SN26, and SN27, which represented the diversity of signals in the single dilution BLI-ISA (Fig 3d). Data from the Total Antibody Binding step were ambiguous, with SP3 slightly separating from SN21 at the 1:2 dilution, although the signal observed in all of the SN samples increased. Data from the Detection step, however, revealed substantial signal improvement of SP3 while maintaining low signal for the SN samples, including SN21, which had produced the most background signal in the single dilution screen.

Overall, our data suggests that BLI-ELISA can detect and rank seropositive samples at a single plasma dilution. In the case of strongly seropositive samples at a 1:8 dilution, the Total Antibody Binding step alone appears sufficient to classify them as seropositive and evaluate antibody levels. While the Detection step provides qualitative confirmation of IgG antibodies at a 1:8 dilution, this step can evaluate antigen-specific IgG levels at a 1:16 dilution. Importantly, the Detection step is able identify all ten SP samples as positive, including moderate and weak seropositive samples, even with a conservative cutoff of the SN mean plus five standard deviations. Similar to ELISA assays, dilution-series BLI-ISA can be used to re-assess ambiguous samples.

### Comparison of RBD versus prefusion Spike in BLI-ISA

To confirm that our observations for the RBD would be applicable more generally, we performed the BLI-ELISA using a trimeric, prefusion-stabilized spike (prefusion Spike) with all the seropositive samples (Fig 4a). Due to sample limitations, we limited our negative controls to the two seronegative samples of which we had the greatest quantity. In general, the Total Antibody Binding step was consistent with what was observed with the RBD. The range in signal between samples was less extreme, which could be due to different loading characteristics of the two antigens, different loading capabilities of the different biosensor surfaces (SA versus HIS1K biosensors), and/or the greater number of epitopes present on the full prefusion Spike compared to the RBD. In addition, the Detection step with the anti-human IgG antibody generally reflected the magnitude in the first step, likely as a result of the lower overall signal.

**Fig. 4.**
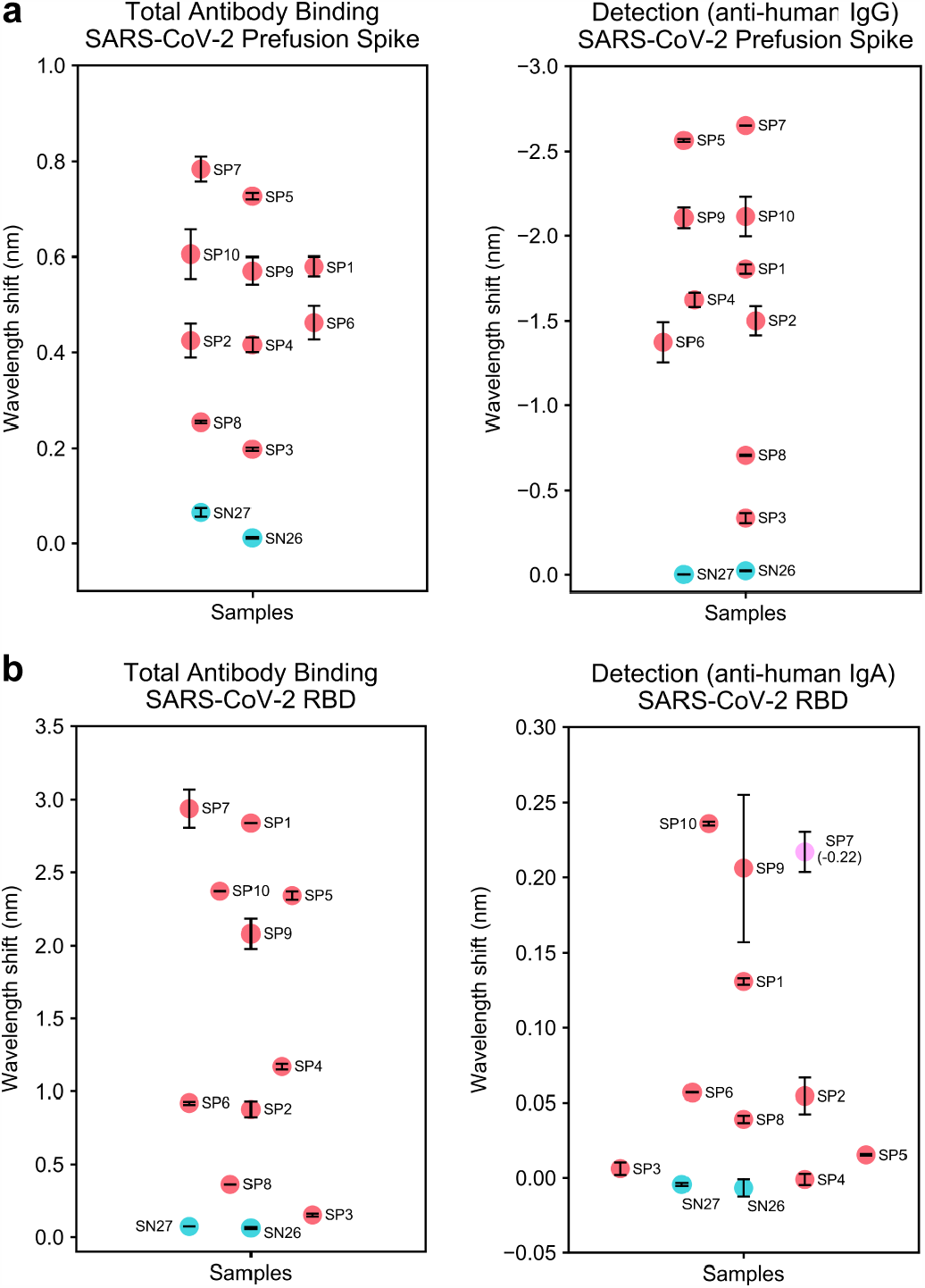
BLI-ISA evaluation of plasma antibodies to SARS-CoV-2 prefusion Spike and plasma IgA to SARS-CoV-2 spike RBD. **a** Single-dilution BLI-ISA to evaluate the presence of prefusion Spike-reactive human antibodies in the pre-pandemic seronegative (SN, cyan) and convalescent seropositive (SP, red) samples. The Total Antibody Binding signal (left) is measured when prefusion Spike-His-loaded HIS1K biosensors are dipped into plasma samples. The Detection signal (right) is measured when prefusion Spike-His-loaded HIS1K biosensors that had been dipped into plasma are subsequently dipped into colloidal gold-conjugated anti-human IgG. **b** Single-dilution BLI-ISA to evaluate the presence of RBD-reactive human antibodies in the samples. The Total Antibody Binding signal (left) is measured when RBD-biotin-loaded SA biosensors are dipped into plasma samples. The Detection signal (right) is measured when RBD-biotin-loaded SA biosensors that had been dipped into plasma are subsequently dipped into colloidal gold-conjugated anti-human IgA. The SP7 dot is colored pink to indicate that this sample had a negative signal (value in parentheses) in the Detection step. All assays were performed with plasma at a 1:8 dilution. Dots represent the mean of biological duplicates, and error bars represent one standard deviation from the mean.

Intriguingly, the Detection step resulted in strong downward, negative curves for the SP samples, while remaining flat for the SN samples (Supplementary Figure 7). The cause for this flip in expected signal is not entirely clear. One possibility is that binding of the anti-human IgG antibody produces a conformational shift or “pull” to antibodies bound to the spike protein, resulting in a change to the sensor surface as revealed by a reduced BLI signal. Alternatively, there may be a higher density of antibodies due to the increased binding sites on the spike, resulting in steric competition with the anti-human IgG antibody that causes weakly bound antibodies to be released. In either case, the signal is specifically triggered by the introduction of the secondary antibody to antigen that has been exposed to SP plasma, suggesting that the qualitative nature of the step remains unimpaired.

### Detecting different antibody isotypes

Although IgG is the most prevalent antibody isotype in circulation, making it ideal for serosurveillance, in some cases it may be desirable to identify different isotypes in a sample or target a specific physiological context where another antibody isotype is most prevalent. IgA, for example, is most predominant in mucosal membranes, but is also present at a low percentage in blood. To assess whether we could specifically detect IgA, we evaluated the signal in the Detection step using a colloidal gold-conjugated anti-IgA antibody (Fig 4b). Not surprisingly, the signals were substantially lower than those observed for the anti-IgG antibody and were undetectable in some cases. However, some samples showed a clear signal, especially several of the strong SP samples. Intriguingly, while the SP samples generally showed a positive signal, SP7 showed a negative curve. This may be consistent with a model of steric competition between weakly bound IgG and the IgA-specific antibody, or due to differences in antibody epitope usage in this individual sample. Excitingly, these results suggest that different isotypes can be detected from a single sample by BLI-ISA, including those that are not the predominant isotype.

## Discussion

Here, we describe BLI-ISA, a novel serological testing method to measure antigen-specific antibodies in plasma utilizing biolayer interferometry. The assay is simple to perform; samples are pipetted into a 96- or 384-well plate, the plate is loaded into the BLI instrument along with single-use biosensors, and the assay program is run, directing the instrument to dip biosensors in different wells for each step (Fig 2 and Supplementary Figure 2). Moreover, the assay is rapid, with real-time data output and full results in <20 minutes following sample preparation. The assay is quantitative and measures both total antibody levels and specific antibody isotypes in the same assay, and single-dilution BLI-ISA signals from diverse seropositive samples align with AUC values obtained by dilution-series ELISA.

Beyond the advantage of short assay times, BLI-ISA has technical advantages over ELISA, IFA, and CLIA in two key ways; (1) BLI-ISA does not require washing of wells or beads, eliminating time-consuming manual washing or use of fluidic instrumentation, and (2) BLI-ISA does not utilize enzyme-based signal amplification (i.e. HRP), which can vary due to differences in temperature, pH, and manufacturing lots of enzyme-conjugated reagent. As a result, our assay overcomes the lab-to-lab variability that can occur with methods that require extensive washing and/or enzyme-based signal amplification. Thus, BLI-ISA provides a solution to standardize other serological testing methods as well as to perform longitudinal studies of biological samples. We do acknowledge a caveat of our BLI-ISA method is the need for BLI instrumentation, however this instrument is becoming more commonplace at research institutions, and in many cases the acquisition of such an instrument would be beneficial due to reduced labor, faster data acquisition time, and reduced data variability compared to other methods. We also acknowledge the relatively small numbers of pre-pandemic samples used for this proof-of-principal study are not sufficient to precisely define the lower limit of detection, however evaluation of more pre-pandemic samples will ultimately make this limit clear.

In a broader sense, BLI-ISA can be adapted, multiplexed, and performed in a high-throughput fashion. Straightforward adaptions can allow for measurement of antibodies against different antigens, as we demonstrated for RBD and prefusion Spike, and/or detection of different antibody isotypes, as we demonstrated for IgG and IgA antibodies. There is also potential to measure antibodies in other biological specimens (e.g. breastmilk, saliva) and also measure other clinically relevant, non-antibody biological molecules in human and animal specimens. Finally, BLI-ISA has the potential to be performed in a high-throughput fashion. While many institutions have BLI instruments that measure 8 or 16 biosensors at a time, the Octet HTX instrument can measure 96 biosensors at a time. In addition, we found that RBD-biotin could be pre-loaded onto SA biosensors with no loss in signal over at least three hours (data not shown), suggesting that biosensors could be pre-loaded to eliminate this step from the assay method and save time. Thus, with antigen pre-loading and the use of an Octet HTX instrument, approximately 3,000 samples could be analyzed in an 8-hour day.

Here, we used BLI-ISA to detect antibodies to SARS-CoV-2 using the spike RBD antigen. We chose this antigen because it is highly selective for antibodies to SARS CoV-2 and because antibodies to the RBD have been shown to correlate with virus neutralization ^8,10,12-17^. We also found that use of prefusion Spike antigen shows similar trends in seroreactivity. Our BLI-ISA method can be immediately implemented for urgent SARS-CoV-2 serological testing needs. First, our method could be used for serosurveillance studies to evaluate seroconversion in communities. Second, and importantly, BLI-ISA can be used to evaluate antibody responses to natural infection and vaccine candidates to define correlates of immunity to SARS-CoV-2 infection. Finally, we believe that BLI-ISA can be developed as a novel diagnostic platform to evaluate antibodies and other biomolecules in clinical specimens, for example to evaluate plasma antibody levels to inform patients on vaccinations, or to quickly identify and prioritize donors for convalescent plasma therapy donation ^37,38^.

## Methods

### Reagents and supplies

Phosphate buffered saline (PBS) tablets (Sigma P4417), Tween-20 (Fisher BP337), dry milk powder (RPI 50488786), ELISA plates (Corning 3590), Goat anti-Human IgG Fc HRP (Thermo Fisher A18817), OPD tablets (Pierce PI34006), bovine serum albumin (BSA) (Fisher BP1600), ChonBlock (Chondrex 9068), Biotinylated SARS-CoV-2 protein RBD His AviTag (Acro Biosystems SPD-C82E9), 4 nm Colloidal Gold-AffiPure Goat Anti-Human IgG Fcg fragment specific (Jackson ImmunoResearch 109-185-098), 4 nm Colloidal Gold-AffiPure Goat Anti-Human Serum IgA alpha chain specific (Jackson ImmunoResearch 109-185-011), Human coronavirus spike glycoprotein Antibody, Rabbit PAb, Antigen Affinity Purified (Sino Biological 40021-T60), SARS-CoV-2 (2019-nCoV) spike Antibody, Rabbit PAb, Antigen Affinity Purified (Sino Biological 40589-T62), SARS-CoV-2 (2019-nCoV) spike Antibody, Rabbit MAb (40150-R007), Anti-SARS-CoV S Therapeutic Antibody (CR3022) (Creative Biolabs MRO-1214LC), Octet Anti-Penta-His (HIS1K) sensor tips (Sartorius ForteBio 18-5120), Octet Streptavidin (SA) sensor tips (Sartorius ForteBio 18-5109), tilted bottom (TW384) microplates (Sartorius ForteBio 18-5080), electroporation cuvettes (MaxCyte SOC4), suspension adapted CHO-S cells (Thermo Fisher R80007). CD-CHO medium (Thermo Fisher 10743029), CD OptiCHO medium (Thermo Fisher 12681011), HisTrap FF (GE Healthcare 17-5286-01), StrepTrap HP (GE Healthcare 28-9075-47), Superdex 200 Increase GL (GE Healthcare 28-9909-44).

### Recombinant SARS-CoV-2 spike proteins

The expression plasmid for the SARS CoV-2 spike RBD-His was obtained from BEI Resources. This pCAGGS plasmid encodes the signal peptide (residues 1-14) and RBD (residues 319-541) of the SARS CoV-2 spike (GenBank: MN908947.3), fused to a C-terminal 6XHis-tag ^12^. To generate the expression plasmid for RBD-biotin, the cDNA encoding the SARS-CoV-2 spike signal peptide, RBD and 6XHis-tag in pCAGGS was sub-cloned by Gibson Assembly into a derivative of pcDNA3.1 ^39^ in frame with a Strep-tag and AviTag at the C-terminus. The pαH expression plasmid encoding the prefusion-stabilized SARS CoV-2 spike trimer was described previously ^32^. Recombinant prefusion-stabilized spike trimer (prefusion Spike-His) produced in ExpiCHO cells was a generous gift from the Almo lab (Albert Einstein College of Medicine)^33^. Recombinant RBD proteins were produced in suspension adapted CHO-S cells. CHO-S cells were maintained in CD-CHO medium supplemented with 8 mM GlutaMAX, 0.1 mM Hypoxanthine, and 0.016 mM thymidine (HT) in shake flasks using a Khuner shaker incubator at 37°C, 8% CO_2_, and 85% humidity. For protein production, CHO-S cells were transfected with purified endotoxin-free DNA using flow electroporation technology (MaxCyte). Transfected cells were grown at 32°C in CD OptiCHO medium supplemented with 2 mM GlutaMAX, HT supplement, 0.1% pluronic, and 1 mM sodium butyrate, supplementing daily with MaxCyte CHO A Feed (comprised of 0.5% Yeastolate, 2.5% CHO-CD Efficient Feed A, 2 g/L Glucose, and 0.25 mM GlutaMAX). On day 8 post-transfection, cells were centrifuged at 4000g for 15 min, and the media was 0.22-μm filtered. For purification of RBD-His, media was diluted with Buffer A (300 mM NaCl, 50 mM NaH_2_PO_4_, 20 mM imidazole [pH 7.4]) and loaded onto a HisTrap column. The column was washed with Buffer A, and RBD-His was eluted with a gradient to Buffer B (300 mM NaCl, 50 mM NaH_2_PO_4_, 225 mM imidazole [pH 7.4]). RBD-His was further purified by size-exclusion chromatography on a Superdex 200 column in PBS and the fractions containing pure monomeric RBD-His were pooled and concentrated to 1.03 mg/ml. For purification of RBD-biotin, media was supplemented with 20 mM Tris pH 8.0 and 150 mM NaCl (TBS), 1 mM EDTA, and BioLock and loaded onto a StrepTrap column. The column was washed with TBS containing 1 mM EDTA, and RBD protein was eluted with a gradient of TBS, 1 mM EDTA, and 2.5 mM desthiobiotin. Additional RBD protein in the media was obtained by dialysis and purification with a HisTrap column as described above. The purest elution fractions from each purification were pooled and dialyzed overnight into PBS. Biotinylation of the AviTag was achieved following published procedures ^40^. Briefly, 46 μM RBD was incubated overnight at room temperature with 3 μM recombinant GST-BirA biotin ligase in PBS containing 5 mM MgCl_2_, 25 mM ATP, and 625 μM D-biotin. RBD-biotin was further purified by size-exclusion chromatography on a Superdex 200 column in PBS and the fractions containing pure monomeric RBD-biotin were pooled and concentrated to 315 μg/ml. All purified recombinant proteins were aliquoted, flash frozen in liquid nitrogen, and stored at -80 °C until use.

### Human samples

Human plasma and serum samples were obtained from several sources. The pre-pandemic seronegative (SN) panel comprised of de-identified samples selected based on the date of collection, before the emergence of SARS-CoV-2. First, human serum samples (n = 25) collected in 2017 were from study participants enrolled in an Institutional Review Board-approved study for development of Lyme disease and other diagnostic tests. To this end 550 samples were collected from individuals on the East Coast and in the Upper Midwest of the United States where Lyme disease is endemic ^41^. Samples were obtained from the Lyme Disease Biobank as part of a research collaboration with Ontera Inc. (Santa Cruz, CA, USA). Second, human plasma samples (n = 2) collected in 2016 were from study participants enrolled in an Institutional Review Board-approved study at UCSC. All participants agreed to sample banking and future research use. The convalescent seropositive (SP) panel comprised 10 de-identified plasma samples from nine individuals, purchased from AllCells (Alameda, CA, USA). To be eligible for plasma donation, prospective donors must have either had COVID-19 symptoms resolved at least 28 days after the diagnosis or suspicion of COVID-19 (i.e. no fever, cough, difficulty breathing, etc.) or been 14 days symptom free with a follow up negative nasopharyngeal/PCR test. Each SP sample had been tested by an Anti-SARS-CoV-2 Total test (Ortho Clinical Diagnostics) and was given a CoV2T score (ranging from 8 to 440) (Table 1). All serum and plasma samples were heated at 56 °C for 1 hour before use.

### ELISA

The ELISA protocol was adapted from a previously published protocol ^12^. ELISA plates (96-well) were incubated overnight at 4 °C with 50 μl per well of 2 μg/ml RBD-His in PBS. After removal of RBD-His, plates were blocked for 1 hour at room temperature with 300 μl per well of 3% non-fat milk in PBS with 0.1% Tween 20 (PBST). After removal of blocking buffer, 100 μl plasma/serum samples diluted 1:50 in 3% milk in PBST were added to wells and incubated for 2 hours at room temperature with gentle shaking. For dilution series ELISAs, plasma/serum samples were first diluted 1:50 in 3% milk in PBST and then diluted 1:4 in series in 3% milk in PBST. The human monoclonal antibody CR3022 antibody, which is reactive to the RBD of both SARS-CoV-1 and SARS-CoV-2, was used as a positive control ^34-36^. The plates were washed three times with PBST. After washing, 100 μl goat anti-human IgG Fc horseradish peroxidase (HRP) conjugated secondary antibody diluted 1:3000 in 1% milk in PBST was added to each well and incubated for 1 hour at room temperature with gentle shaking. Plates were again washed three times with PBST. The plates were washed three times with PBST, followed by addition of 100 μl o-phenylenediamine dihydrochloride (OPD) solution to each well. The substrate was left on the plates for exactly 10 minutes and then the reaction was stopped by adding 50 μl per well of 3 M hydrochloric acid. The optical density at 490 nm (OD490) was measured using a Molecular Devices Spectramax plate reader. The background value was set at an OD490 of 0.051 based on the average PBS measurement and subtracted from all data prior to curve fitting and AUC calculations. The AUC values were calculated by fitting a four-parameter logistic regression model to the OD490 data of each sample using the curve fit algorithm from the SciPy Python Library ^42^ followed by integration between the upper and lower bounds of the data.

### BioLayer Interferometry ImmunoSorbent Assay (*BLI-ISA*)

BLI-ISA studies were performed on an Octet RED384 instrument at 24 °C with shaking at 1000 rpm. BLI assay buffer consists of 2% BSA in PBST, which was 0.22-μm filtered. Before use, anti-penta-His (HIS1K) or (anti-biotin) streptavidin (SA) biosensors (were loaded into the columns of a biosensor holding plate and pre-hydrated in BLI assay buffer for 10 minutes. Tilted bottom 384-well microplates were loaded with 45 μl per well. The assay plate was prepared as follows: column 1 (BLI assay buffer), column 2 (2-12 μg/ml RBD or prefusion Spike in BLI assay buffer), column 3 (25% ChonBlock in BLI assay buffer), column 4 (plasma/serum samples diluted 1:8 in 25% ChonBlock in BLI assay buffer), column 5 (BLI assay buffer), and column 6 (4 nm Colloidal Gold-AffiPure Goat Anti-Human IgG or IgA secondary antibody diluted 1:10 in BLI assay buffer). For dilution series studies, samples were diluted into stock solutions of ChonBlock in BLI assay buffer to yield a 25% ChonBlock solution after plasma dilution. RBD-biotin purchased from Acro Biosystems was used at 2 μg/ml for 1:8 single-dilution studies.

RBD-biotin produced in-house at UCSC was not fully biotinylated and required use at 10 μg/ml to achieve the same loading signal. RBD-biotin produced at UCSC was used for dilution-series studies and anti-human IgA studies. RBD-His and Prefusion Spike-His were used at 10 μg/ml.

The BLI-ISA method was set as follows. Baseline1 (60 sec) in column 1 (Equilibration), Loading (120 sec or 600 sec) in column 2 (Antigen Loading: RBD or prefusion Spike, respectively), Baseline2 (60 sec) in column 3 (Wash), Association1 (600 sec) in column 4 (Total Antibody Binding), Baseline3 (60 sec) in column 5 (Wash), and Association2 (180 sec) in column 6 (Detection: anti-human IgG or IgA). Loading of RBD-biotin over 120 seconds onto SA sensor tips resulted in a wavelength shift signal of ∼2.2 nm (Supplementary Figure 4). Loading of RBD-His or prefusion Spike-His over 600 seconds resulted in a wavelength shift signal of ∼1.0 nm (Supplementary Figure 4). We note that loading density on sensor tips had little effect on antibody binding signals in Association 1 or 2.

A Python program was written to automate analysis of BLI-ISA data. Raw data .csv files were exported from the Octet Data Analysis software and read by our program. Our script determined the Total Antibody Binding (Association 1) value by subtracting the average wavelength shift of seconds 2-4 of this step from the average shift of the last 5 seconds of this step. Similarly, the Detection (anti-human IgG or IgA binding) (Association 2) value was determined by subtracting the average of the data 1-2 seconds after the start of this step from the average of the data from the last 5 seconds of this step. Complete raw data traces were also plotted and inspected to ensure proper antigen loading onto sensor tips where applicable. This Python program named bli_plotter is available for adaption to other BLI-ISA studies and has been deposited on GitHub (https://github.com/nlorigroach/bli_plotter).

## Data Availability

The raw data for all data referred to in the manuscript is available upon request.

## Acknowledgements

The authors are grateful for the support from the following individuals. Natalia Herrera, Nicholas Morano, Scott Garforth, and Steven Alamo (Albert Einstein College of Medicine) provided recombinant prefusion Spike protein. The Lyme Disease Biobank donated pre-pandemic human serum samples via a research collaboration with Ontera Inc. Phillip Berman shared a derivative of pcDNA3.1 plasmid and allowed use of MaxCyte electroporation instrumentation and Kuhner shaker incubators. The following reagent was produced under HHSN272201400008C and obtained through BEI Resources, NIAID, NIH: Vector pCAGGS Containing the SARS-Related Coronavirus 2, Wuhan-Hu-1 Spike Glycoprotein Receptor Binding Domain (RBD), NR-52309. We thank UCSC colleagues for advice, especially Susan Carpenter, Camilla Forsberg, Ed Green, David Haussler, Sofie Salama, Beth Shapiro, and Josh Stuart.

Funding for this research came from the University of California Office of the President, the UC Santa Cruz Office of Research, the Michael-David Family Foundation, and donors to the UC Santa Cruz Respiratory Virus Fund. Funding for the purchase of the Octet RED384 instrument was supported by the NIH S10 shared instrumentation grant 1S10OD027012-01.

## Author Contributions

J.V.D., N.L.R., and R.M.D. conceived the study. S.M.O. expressed RBD proteins in CHO cells. J.V.D. and R.M.D. purified RBD-His and RBD-biotin proteins and acquired data. N.L.R. wrote the BLI-plotter program. J.V.D., N.L.R., and R.M.D. analyzed the data. D.L.A. and J.M.K. contributed unique reagents and consulted on experimental design. J.V.D. and N.L.R. prepared figures. J.V.D. and R.M.D. write the manuscript. All authors edited the manuscript.

